# Defective monocyte enzymatic function and an inhibitory immune phenotype in HIV-exposed uninfected African infants in the era of anti-retroviral therapy

**DOI:** 10.1101/2021.07.26.21261143

**Authors:** Louise Afran, Kondwani C. Jambo, Wilfred Nedi, David JC Miles, Anmol Kiran, Dominic H Banda, Ralph Kamg’ona, Dumizulu Tembo, Annette Burger, Eleni Nastouli, Brigit Ferne, Henry C Mwandumba, Paul Moss, David Goldblatt, Sarah Rowland-Jones, Adam Finn, Robert S Heyderman

**Author notes:** Correspondence to: Louise Afran, Malawi-Liverpool-Wellcome Trust Clinical Research Programme, P.O Box 30096, Chichiri, Blantyre 3, Malawi., or Robert S Heyderman, Division of Infection and Immunity University College London, Rayne Building, 5 University Street, London, WC1E 6JF.

## Abstract

HIV-Exposed Uninfected (HEU) infants are a rapidly expanding population in sub-Saharan Africa and are highly susceptible to disease caused by encapsulated bacteria in the first year of life. The mechanism of this increased risk is still poorly understood. We therefore investigated if HIV exposure dysregulates HEU infant immunity and if this is amplified by human herpes virus infection (HHV). Here, we compared monocyte enzymatic function, innate and adaptive immune cell phenotype, and vaccine-induced antibody responses between HEU and HUU infants. We demonstrate altered monocyte phagosomal function and B cell subset homeostasis, and lower vaccine-induced anti-*Haemophilus influenzae type b* (*Hib)* and anti-Tetanus Toxoid (TT) IgG titers in HEU compared to HUU infants. There was no difference in the prevalence of HHV infection between HEU and HUU infants. Our findings suggest that even in the era of antiretroviral therapy (ART)-mediated viral suppression, HIV exposure dysregulates monocyte and B cell function during a vulnerable period of immune maturation in infancy. This may contribute to the high rates of invasive bacterial disease and pneumonia in HEU infants.

## Introduction

HIV-exposed uninfected (HEU) infants are more susceptible to infection-related morbidity and mortality ^1, 2^. HEU infants are particularly vulnerable to invasive bacterial disease^3–7^ particularly, pneumonia^8–13^ and diarrhea^14, 15^, they have more frequent hospitalisations, more severe infections and increased risk of treatment failure. However, the mechanism of this increased vulnerability remains unknown. The global population of children who are HEU is substantial, estimated at 1.2 million births annually, mainly within developing countries^16^. A coordinated strategy is therefore necessary to ensure their optimal health and wellbeing^17^.

The vulnerability of HEU infants is a likely complex intersection of HIV-exposure immune profile ‘remodelling’, an ‘inflammatory’ maternal cytokine milieu^18^, time of antiretroviral therapy (ART) initiation^19^, ART use^20^ and prophylactics ^21^, increased exposure to maternal viral and bacterial pathogens, and host microbial and environmental factors. Together these result in a more permissive state for the development of infections ^1^. Many observational studies have reported immunological abnormalities in HEU infants ^22^ including highly differentiated T-cells ^23–29^ and B-cell subsets ^30^, altered responses to vaccines ^31–33^, functional impairment of natural killer cells ^34^ and monocytes ^35–37^. Furthermore, few studies have considered early transmission of immunomodulatory human herpes virus’ (HHVs), cytomegalovirus (CMV)^38^ and Epstein Barr virus (EBV)^39^ recrudescence during pregnancy on HEU immunity. HHVs are important as they are implicated in delayed growth^40, 41^, poor neurodevelopment, altered mitochondria DNA^42^ and inflammation in HEU infants^43, 44^.

Due to the successful HIV test and treat strategy globally, the prevalence of individuals receiving ART has risen considerably^16^. Consequently, the number of HEU infants born to mothers receiving ART has increased markedly ^45, 46^, but despite expanded maternal use of ART ^47^ or implementation of prevention-of-mother-to-child-transmission (PMTCT) programmes ^48^, the risk of infection-related morbidity and mortality among HEU infants remains high^2^, particularly the risk of encapsulated bacterial infection^7, 9, 49–51^. The immune profile amongst HEU infants in this context is not well documented. We have therefore addressed the question of whether HIV exposure and HHVs dysregulate infant immunity and/or response to primary vaccination. By comprehensive cellular and serological assessment, we show impaired monocyte phagosomal function, altered B cell homeostasis and selective impairment of vaccine-induced anti-Hib antibody response in HEU babies less than 6 months of age. These findings suggest acquired defects in innate and adaptive immunity, heightened regulation of HEU immune responses, which may increase susceptibility to invasive bacterial disease and pneumonia.

## Results

### Participant characteristics

We recruited two cohorts to evaluate the impact of HIV exposure and herpes virus infection on immunity at birth and in early infancy. A birth cohort comprised 34 HIV-infected and 44 HIV-uninfected pregnant women. A longitudinal infant cohort comprised 43 HIV-infected and 61 HIV-uninfected mother-infant pairs, who were followed at 3 timepoints, 5-9, 14-15 and 18-23 weeks of age, corresponding to the Malawian routine infant vaccine schedule of: BCG at birth, then pentavalent vaccine at 6, 10 and 14 weeks.

In the birth cohort, two babies were excluded from the analysis due to death and HIV-positivity detected by digital droplet PCR. HIV-infected pregnant women received ART (Option B+) for an average of 18.7 (range 1-143) months, with a mean nadir CD4+ T-cell count of 294 (range 8-892) and were more likely to have had an elective caesarean birth, compared to HIV-uninfected pregnant women (Table 1).

**Table 1.** Participant characteristics among HEU new-borns, infants and HUU controls.

|  | New-born Birth cohort |  |  | Longitudinal Infant cohort |  |  |
| --- | --- | --- | --- | --- | --- | --- |
|  | HIV- | HIV+ | *p-value* | HIV- | HIV+ | *p-value* |
| **<sup>a</sup>Mothers age in years (IQR)** | 27.6 (23.6-32.95) | 29.7(25.95-32.2) | 0.86 | 21.9 (19.5-26.2) | 28.8 (25.2-33.1) | 0.37 |
| **<sup>a</sup>Mothers ART no.(%)** | N/A | 34/34 (100) | - | N/A | 37/43 (86) | - |
| **<sup>a</sup>Mothers time on ART months (range)** | N/A | 18.17 (1-143) | - | N/A | 9.28 (0-72) | - |
| **<sup>a</sup>Mothers CD4+ cells μL median (IQR)** | N/A | 294 (8-892) | N/A | N/A | 409 (159-823)* | - |
| **<sup>a</sup>Mothers no. caesarean Section (%)** | 0/44 (0) | 14/34 (39) | **0.01** | - | - | - |
|  | HUU new-born | HEU new-born | *p-value* | HUU infant | HEU infant | *p-value* |
| **<sup>a</sup>Female Sex of child** | 22/44 | 15/34 | 0.65 | 29/61 (55) | 22/43 (39) | 0.24 |
| **HIV DNA PCR test result of child at birth** | - | 31/34 (100) |  | - | 43/43 | - |
| **HIV DNA PCR test result of child at 6 weeks** | - | 34/34 (100) |  | - | 36/43 (83.7)* | - |

Abbreviations: IQR, interquartile range; No., number; %=percentage; μL= microliter; dPCR=digital PCR, RDT = rapid diagnostic test performed on whole blood. <sup>a</sup> Calculated using Mann-Whitney for continuous variables and Fishers exact test for categorical variables.
<sup>b</sup>Result indeterminant. \*7/43 unconfirmed HIV status, \*\*3 mothers had missing CD4 data, \*\*\*self-reported exclusive breastfeeding.

In the longitudinal infant cohort, HIV-infected mothers had received option B+ for an average of 9.28 (range 1-72) months at the time of enrollment and had a mean nadir CD4+ T-cell count of 409 (range 159-823). There was no difference between maternal age or breastfeeding status but mode of delivery was more often caesarean section in HIV-infected mothers compared to HIV-negative mothers (Table 1).

### Impaired CD14^+^ monocyte function in HEU new-borns

Monocytes are an important first line of defence against invasive bacteria, particularly in early life, before the maturation of adaptive immunity ^52, 53^ and are crucial in controlling bacterial pneumonia^54–56^. We used a flow cytometry-based whole blood reporter bead assay^57^ to assess monocyte function in cord blood (Figure 1a-b; Supplementary Figure 1). First, we assessed the ability of monocytes to internalise Alexa Fluor 405-labeled IgG-coated reporter beads at 1 hour post co-incubation, as a proxy of uptake capacity. We showed that the proportion of monocytes that internalised reporter beads was similar between HEU infants and HUU controls (Figure 1c). Second, we assessed the phagosomal superoxide burst activity, an important component of intracellular killing by phagocytes ^58^, and found that phagosomal superoxide burst activity was lower in monocytes from HEU infants compared to HUU controls (Figure 1d). Third, we assessed the phagosomal bulk proteolytic activity, an important intracellular protein degradation mechanism that impacts antigen-presentation ^59^, and showed that the phagosomal bulk proteolytic activity was lower in monocytes from HEU infants compared to HUU controls (Figure 1e). Collectively, these data indicate altered monocyte phagosomal functional capacity at birth in HEU new-borns.

**Figure 1.**
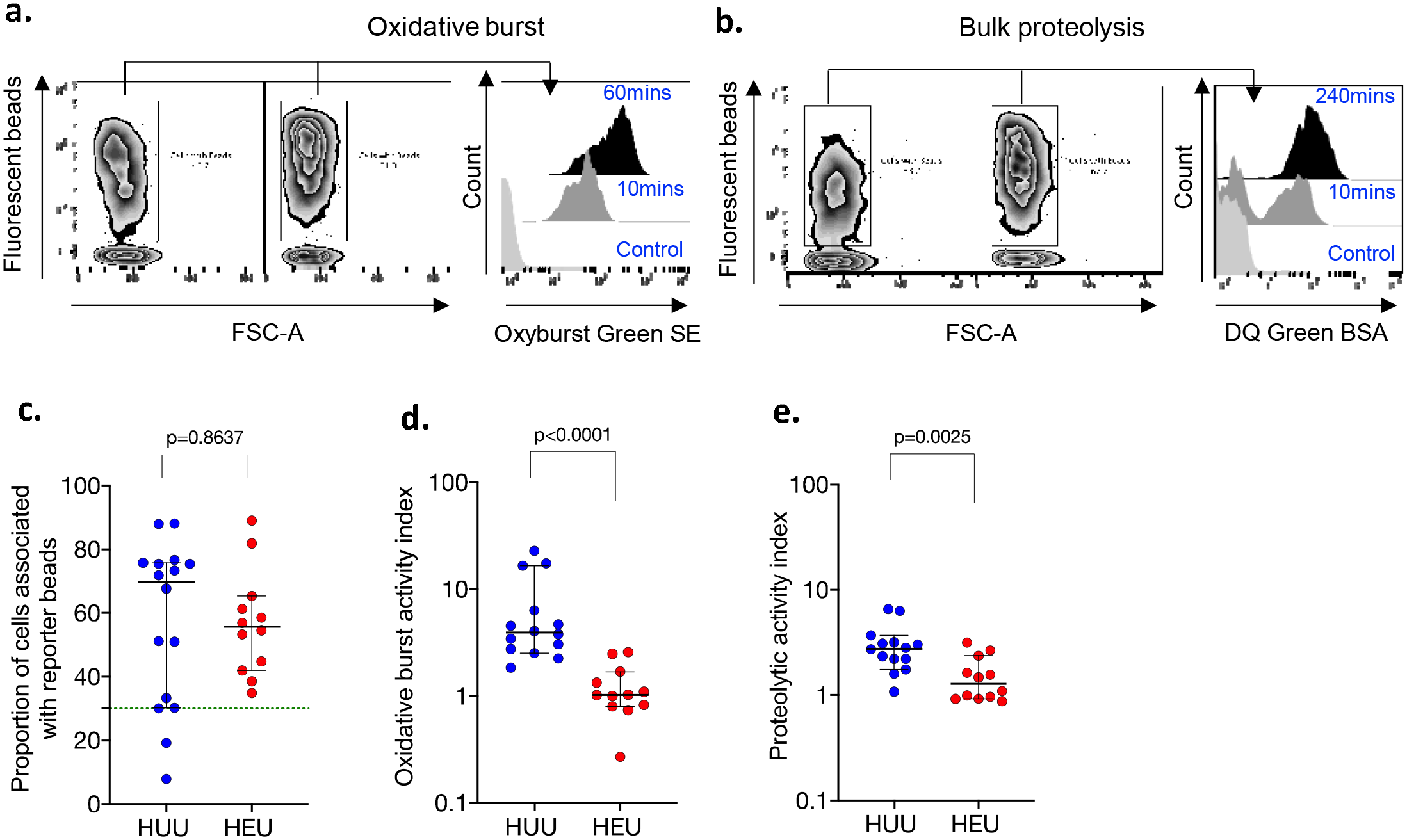
Monocyte phagosomal functional capacity in HEU new-borns and HUU controls. The proportion of CD14+ cells that performed **a)** phagosomal superoxide burst activity, **b)** phagosomal bulk proteolytic activity, **c)** and that were associated with beads, **d)** the phagosomal superoxide burst activity index and **e)** the phagosomal bulk proteolytic activity index. The readout for the assay are reported as the median fluorescent intensity of the reporter fluorochrome at 60mins:10mins and 240mins:10mins for oxidative burst and bulk proteolysis, respectively. The activity index was calculated using a ratio of the reporter fluorochrome over the the calibration fluorochrome. Only individuals with an uptake of greater than ≥30% were used in the phagosomal analysis. Data are presented as medians and analysed using Mann Whitney U test (HUU, n=16; HEU, n=12).

### Inhibitory B cell phenotype and reduced T cell mitogen response in HEU new-borns

Having demonstrated altered monocyte phagosomal function which is part of innate immunity in HEU new-borns, we next sought to investigate whether B and T cell subsets are dysregulated. In the B cell compartment, we observed similar distributions of B cell subsets between HEU and HUU new-born babies, including, CD10^-^CD21^+^CD27^-^ (naive), CD10^-^ CD21^-^CD27^+^ (resting memory), CD10^-^CD21^+^CD27^+^ (activated memory), CD10^-^CD21^-^ CD27^-^ (tissue-like memory) and CD10^+^CD27^-^ (immature transitional) B cells (Figure 2a). However, we found a higher proportion of Fc-receptor like 4 (FcRL4^+^) expressing B cells in HEU new-borns, but no statistically significant differences in the proportion of PD-1 or CD95-expressing B cells, when compared to HUU controls (Figure 2b). FcLR4^+^ inhibits B cell activation through the B cell receptor (BCR) and is a marker of B cell exhaustion in chronically HIV-infected adults ^60^, which suggests increased B cell regulation in HEU new-born babies^61^.

**Figure 2:**
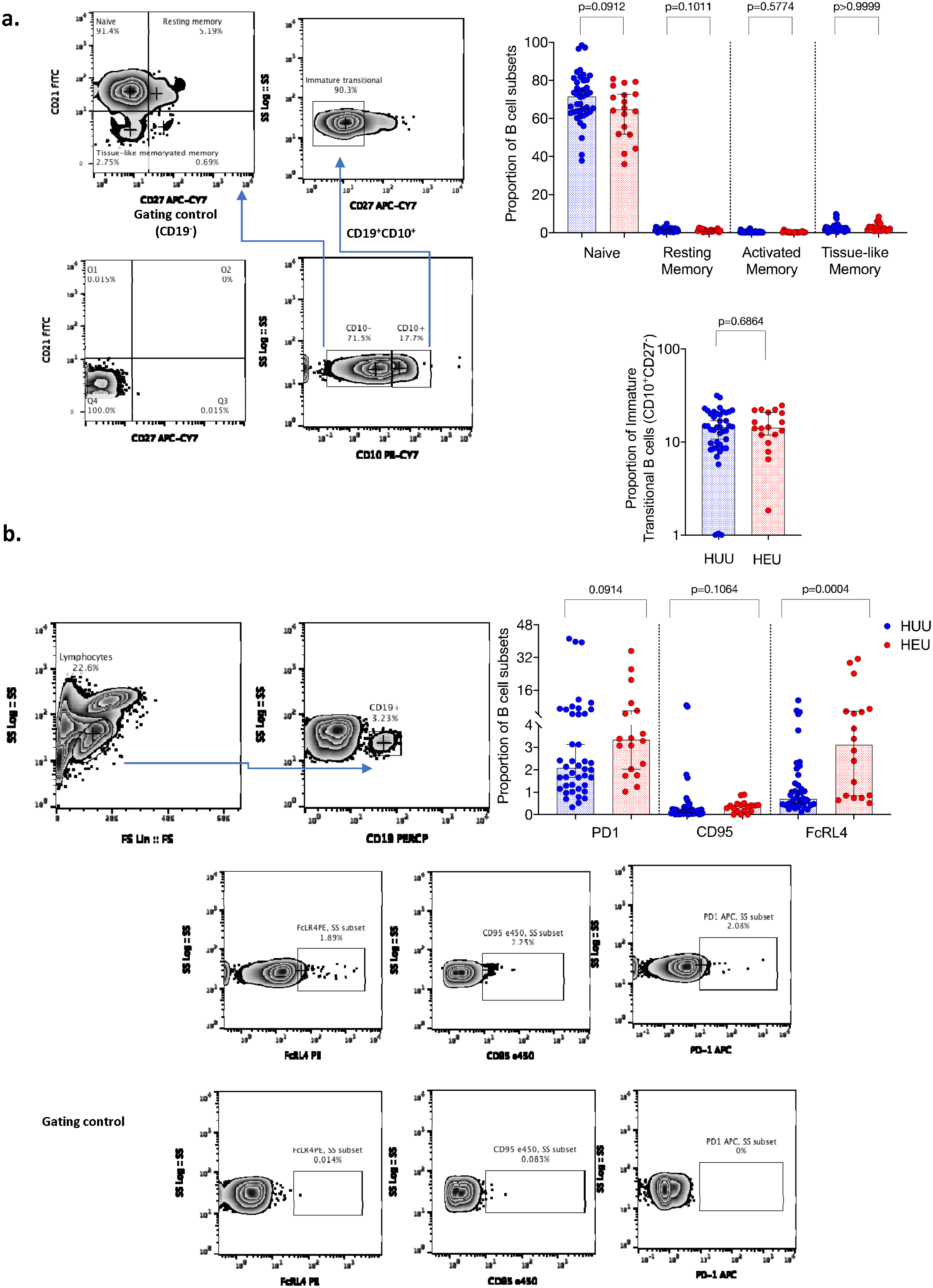
Characterisation of B cell immune profiles in HEU and HUU new-borns. Cord blood was stained with the following fluorochrome-conjugated antibodies, anti-CD19 APC, anti-CD10 PE-Cy7, anti-CD21-FITC and anti-CD27 APC-CY7. Singlets were definedusing FSC-A vs. FSC-H parameters and lymphocytes were gated using SSC-A and FSC-A. B cells were then gated using CD19 against SSC-A. **a)** The proportion of B cell subsets were clasified using CD10, CD21 and CD27 as follows, **b)** CD10^-^CD21^+^CD27^-^ (naive), CD 10^-^ CD21^-^CD27^+^ (resting memory), CD10^-^CD21^+^CD27^+^ (activated memory), CD10^-^CD21^-^ CD27^-^ (tissue-like memory) and **c)** CD10^+^CD27^-^ (immature transitional). **d-e)** Expression of CD95e450, FcLR4 PE and PD-1 APC (exhausted and activatory inhibited B-cells) were gated as a proportion of CD19+ cells. Data are presented as medians [IQR] and analysed using Mann Whitney U test (HUU n=42, HEU n=18).

In the T cell compartment, we found that the proportion of naïve and memory, CD4+ and CD8+ T cell subsets were similar between HEU new-borns and HUU controls (Figure 3a-b). There were also no differences between the two groups in the proportion of CD57 or PD-1-expressing CD4^+^ and CD8^+^ T cells (Figure 3c-d), which are classical markers of T cell immune senescence and exhaustion in chronic HIV infection^62, 63^. IFNγ production in response to tuberculin purified protein derivative (PPD) in an 18-hour ELISpot assay was similar in HEU new-borns compared to controls. In this population BCG is received soon after birth. However, IFNγ production in response to the selective T cell mitogen phytohemagglutinin (PHA)^64^ was reduced in HEU new-borns compared to controls (figure 3e), indicating selective regulation of T-cell responses.

**Figure 3.**
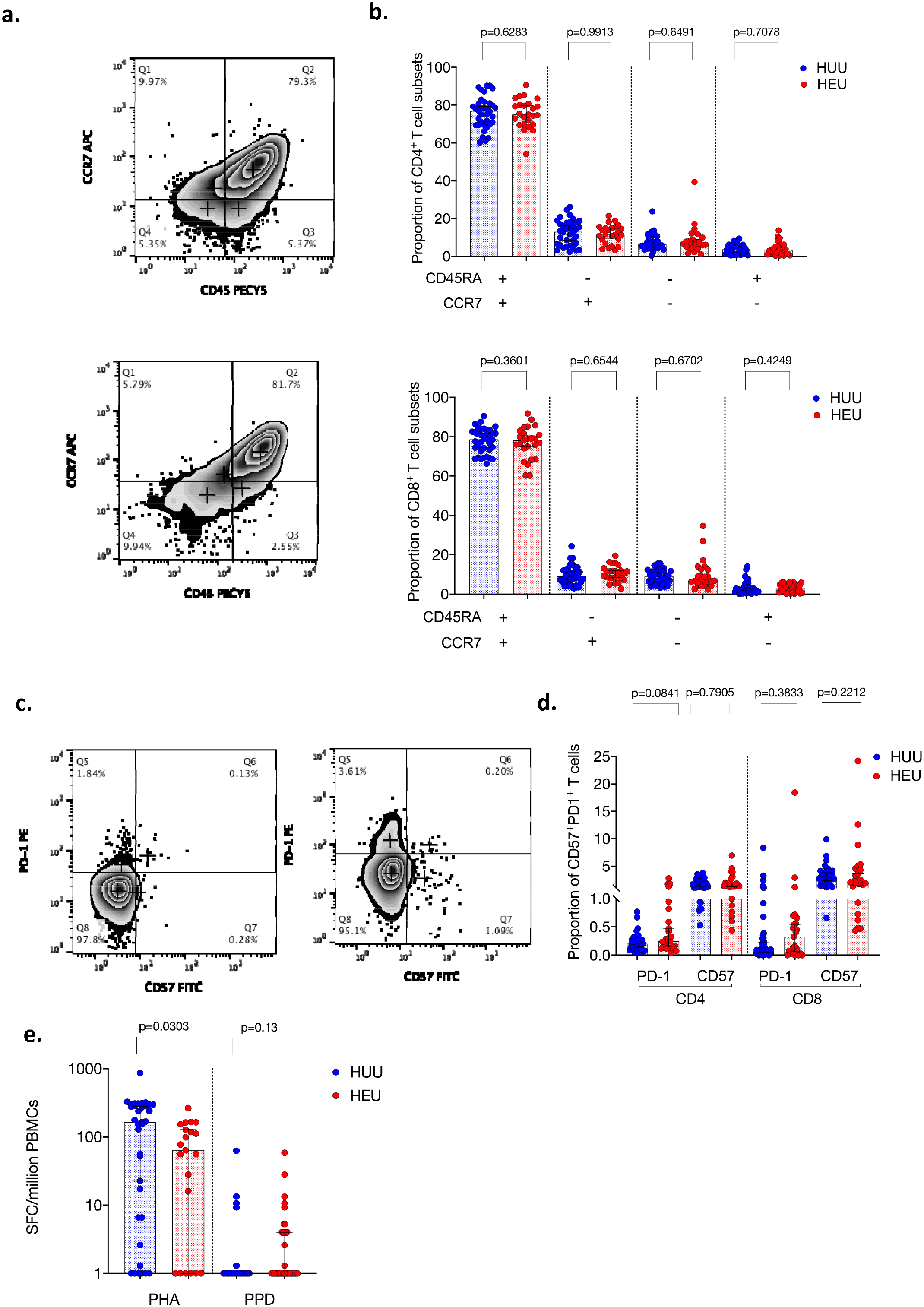
Characterisation of T cell subsets in HEU and HUU new-borns. **a)** Whole cord blood was stained with the following fluorochrome-conjugated antibodies, anti-CD3 APCCY7, anti-CD4 Pacific Blue, anti-CD8-FITC, anti-CCR7 APC and anti-CD45RA PECY7. Singlets were defined using FSC-A vs. FSC-H parameters and lymphocytes were gated using SSC-A and FSC-A. T cells were then gated using CD3 against SSC-A, then a CD4 versus CD8 plot was used to separate the two main T cell subsets. **b)**CD4^+^ and CD8^+^ T cell subsets were clasified using CCR7 and CD45RA as follows CCR7-CD45RA-(effector memory), CCR7^+^CD45RA-(central memory), CCR7^+^CD45RA^+^ (naïve) and CCR7^-^ CD45RA^+^(terminally-differentiated). **c)** Whole cord blood was stained with the following fluorochrome-conjugated antibodies, anti-CD3 APCCY7, anti-CD4 Pacific Blue, anti-CD8-PECY7 and CD57 FITC, PD1-PE. Singlets were defined using FSC-A vs. FSC-H parameters and lymphocytes were gated using SSC-A and FSC-A. T cells were then gated using CD3 against SSC-A, then a CD4 versus CD8 plot was used to separate the two main T cell subsets. **d)** CD4^+^ and CD8^+^ T cell subsets were clasified using CD57 (senescent) and PD-1 (exhausted). (HUU, n =36; HEU, n =25). **e)** Isolated CBMCs were incubated with PHA or PPD for 18 hours and IFN-γ producing cells were detected on a 96-well microtitre ELISpot plate. The frequency of SFCs/million CBMCs are plotted for all subjects. (HUU, n =34; HEU, n =22) Data analysed using Fisher’s exact test.

### Persistent dysregulation of the B cell compartment in HEU infants

Having found an altered innate compartment and increased regulatory B cell phenotype at birth, we then investigated the impact of HIV exposure on adaptive immunity in the first few months of life in the longitudinal infant cohort. We found that the proportions of immature transitional and tissue-like memory B cells were lower in HEU infants than HUU controls (Figure 4a), two B cell subsets that are selectively dysregulated in HIV infection ^65, 66^. However, as seen in the new-born cohort, the proportions of naïve and memory, CD4+ and CD8+ T cell subsets, were similar between HEU infants and HUU controls (Figure 4a-b).

**Figure 4:**
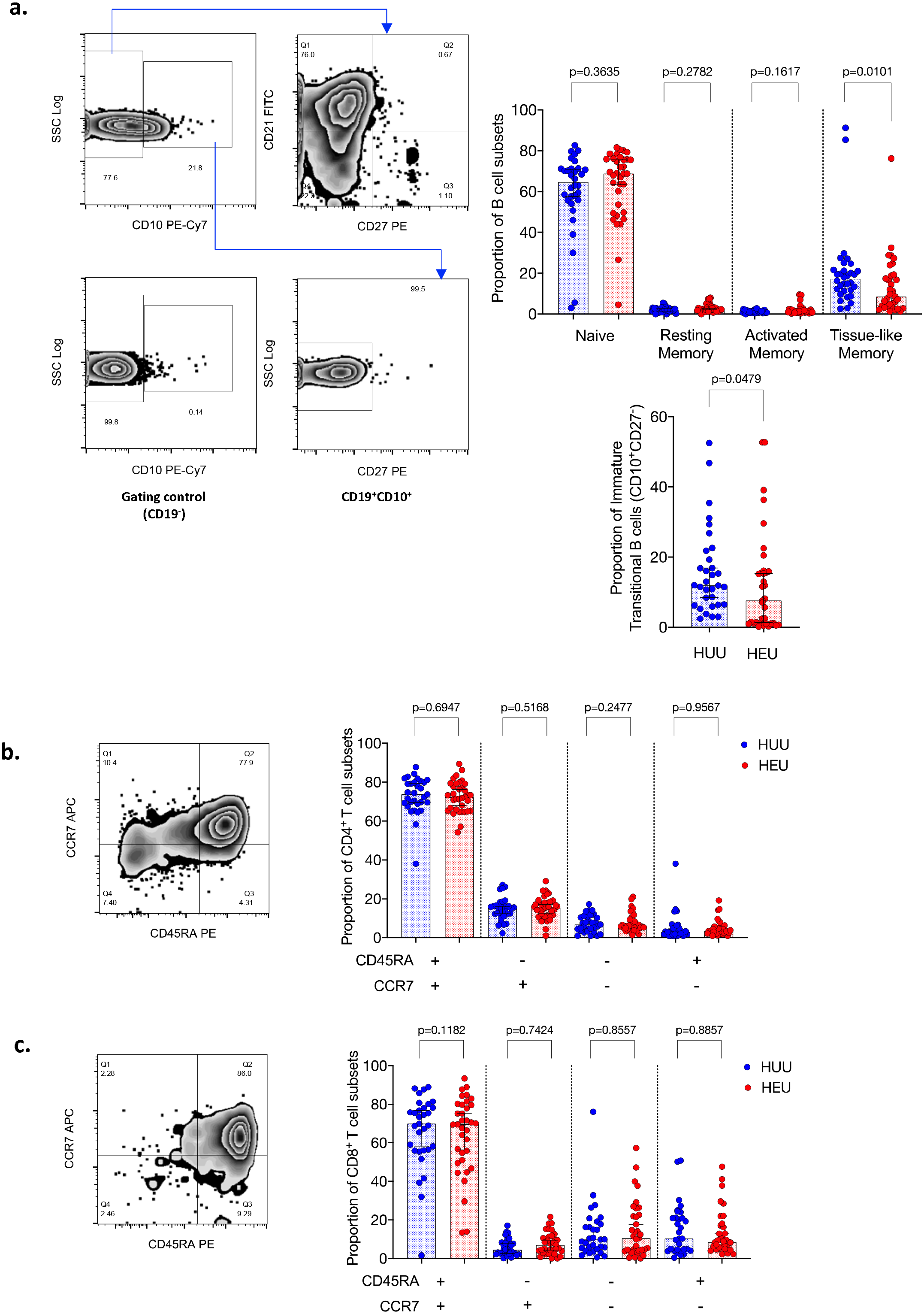
Characterisation of B cell and T cell subsets in HEU and HUU infants. Cord blood was stained with the following fluorochrome-conjugated antibodies, anti-CD19 APC, anti-CD10 PE-Cy7, anti-CD21-FITC and anti-CD27 PE. Singlets were defined using FSC-A vs. FSC-H parameters and lymphocytes were gated using SSC-A and FSC-A. B cells were then gated using CD19 against SSC-A. **a)** The proportion of B cell subsets were clasified using CD10, CD21 and CD27 as follows, **b)** CD10^-^CD21^+^CD27^-^ (naive), CD10^-^ CD21^-^CD27^+^ (resting memory), CD10^-^CD21^+^CD27^+^ (activated memory), CD10^-^CD21^-^ CD27^-^ (tissue-like memory) and **c)** CD10^+^CD27^-^ (immature transitional). Whole blood was stained with the following fluorochrome-conjugated antibodies, anti-CD3 APCH7, anti-CD4 Pacific Blue, anti-CD8-FITC, anti-CCR7 APC and anti-CD45RA PE. Singlets were defined using FSC-A vs. FSC-H parameters and lymphocytes were gated using SSC-A and FSC-A. T cells were then gated using CD3 against SSC-A, then a CD4 versus CD8 plot was used to separate the two main T cell subsets. **d-e)**CD4^+^ and CD8^+^ T cell subsets were clasified using CCR7 and CD45RA as follows CCR7-CD45RA-(effector memory), CCR7^+^CD45RA-(central memory), CCR7^+^CD45RA^+^ (naïve) and CCR7^-^ CD45RA^+^(terminally-differentiated). Data are presented as medians [IQR] and analysed using Mann Whitney U test (HUU n=42, HEU n=31).

### Increased IFNγ production to PPD in a T-cell ELISpot assay in HEU infants

To determine if T-cell responses were altered in HEU infants, we evaluated IFNγ spot forming cells (SFCs) to tetanus toxoid (TT), hepatitis B (Hb) and purified protein derivative (PPD) in an 18-hour T-cell ELIspot in infants aged 5-9 weeks of age. We observed increased IFNγ SFCs/million PBMCs to PPD amongst HEU infants compared to HUU controls, but similar Hb, TT and PHA responses between the two groups (Supplementary figure 5a). These data indicate that HEU infants’ antigen-specific responses to PPD following BCG vaccination at birth have the potential to be greater in capacity than HUU infants.

### Impaired IgG antibody responses to routine pentavalent vaccination in HEU infants

In the longitudinal infant cohort, having demonstrated B-cell dysregulation, we interrogated vaccine-induced memory B cell antibody responses to polysaccharide and protein antigens that are in the Malawian infant primary vaccination series. This included PCV13 and Pentavalent Vaccine (DPT-HepB-Hib) which protect against several important infant disease-causing bacteria, we focused on: *Streptococcus pneumoniae* (13 serotypes) and *Haemophilus influenza* type b (Hib), *Clostridium tetani* (tetanus toxoid) and *Corynebacterium diphtheriae* (diphtheria toxoid).

Following 3 vaccine doses, we found lower anti-*Hib* and anti-diphtheria toxoid (DT) titers, but similar anti-TT titers in HEU infants, compared to HUU controls (Figure 5a-c). In the mothers of these infants, anti-*Hib* IgG and anti-TT IgG titres but not anti-DT titers were lower in HIV-infected mothers than HIV-uninfected controls (Figure 5d-f). Having shown the B cell compartment produces differential antigen specific immunoglobulins to vaccine antigens, we next tested whether HIV exposure influences the levels of vaccine-induced functional antibody. Using a Multiplex Opsonophagocytosis Assay (MOPA), we measured opsonophagocytic activity of 13 vaccine serotypes from the PCV13 (1, 3, 4, 5, 6A, 6B, 7F, 9V, 14, 18C, 19A, 19F, 23F) in infant sera. We found no difference in the geometric mean opsonophagocytic index (GMOI) of 13 pneumococcal serotypes and geometric mean concentration (GMC) of serotype-specific IgG titres, between HEU infants and HUU controls (Table 2).

**Figure 5.**
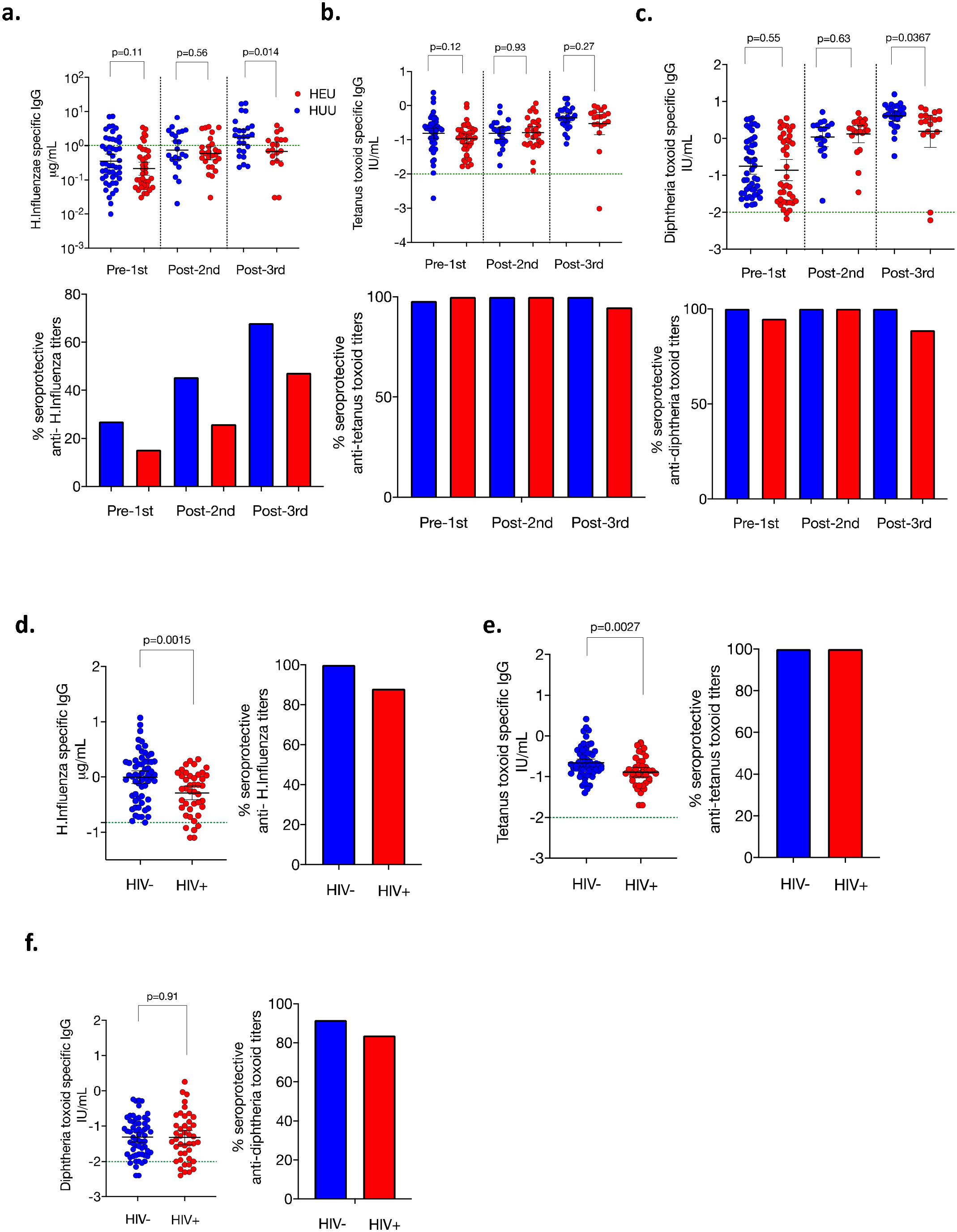
Infant and maternal antibody responses. Preceding and following Penta-DTwPHibHepB vaccination we measured vaccine titers using an ELISA to **a**) anti-Hib IgG **b**) anti-TT IgG **c**) anti-DT IgG in infant serum at 5-7 (HUU, n=50; HEU n=39), 14-15 (HUU, n=22; HEU, n=27) and 18-23 (HUU, n=25; HEU, n=19) weeks. HIV-uninfected (n=61) and HIV-infected (n=43) and maternal titres for **d**) anti-*Hib* IgG **e**) anti-TT IgG **f**) anti-DT IgG are depicted. Blue circles are controls and red are HEU infants or HIV-infected mothers. Green dotted horizontal line represents cut-off for protective titers. Data are presented as medians and analysed using Mann Whitney U test. Minimum putative protective titres are 0.15ug/mL (passive) and 1.0ug/mL (acquired) for *Hib* ^124^, and 0.01IU/mL for TT ^125^and DT _126._

**Table 2.** Robust opsonising and killing function of anti-pneumococcal capsular polysaccharide specific IgG antibodies in infant serum.

| PCV13 Serotypes | HUU infants GMOI (95% CI) | HEU infants GMOI (95% CI) | *p-value* | HUU infants GMC (95% CI) | HEU infants GMC (95% CI) | *p-value* |
| --- | --- | --- | --- | --- | --- | --- |
| 4 | 52.70<br>(12.02 – 231.1) | 69.57<br>(8.32 – 581.9) | 0.95 | 0.52<br>(0.19-1.42) | 0.9<br>(0.2-4.03) | 0.66 |
| 6B | 39.97<br>(7.27 – 219.6) | 65.58<br>(5.72 – 752.1) | 0.76 | 0.39<br>(0.2-0.76) | 0.55<br>(0.17-1.83) | 0.75 |
| 14 | 169<br>(37.31 – 765.2) | 223<br>(32.02 - 1553) | 0.82 | 0.46<br>(0.2-1.08) | 0.84<br>(0.21-3.3) | 0.48 |
| 23F | 48.82<br>(10.19 – 233.9) | 78.70<br>(7.61 – 813.5) | 0.80 | 0.38<br>(0.18-0.79) | 0.51<br>(0.15-1.77) | 0.99 |
| 6A | 46.37<br>(5.9 – 364.3) | 74.07<br>(5.68 – 966.9) | 0.72 | 0.29<br>(0.16-0.52) | 0.51<br>(0.15-1.8) | 0.32 |
| 9V | 65.79<br>(11.18 – 387.1) | 82.85<br>(9.02 – 761.2) | 0.93 | 0.4<br>(0.17-0.91) | 0.74<br>(0.19-2.9) | 0.42 |
| 18C | 55.35<br>(11.88 - 258) | 70.12<br>(8.6 – 570.9) | 0.80 | 0.71<br>(0.24-2.07) | 1.17<br>(0.18-7.48) | 0.99 |
| 19F | 74.04<br>(14.81-370.2) | 92.55<br>(11.48-746) | 0.93 | 0.65<br>(0.3-1.42) | 1.08<br>(0.31-3.81) | 0.49 |
| 1 | 11.22<br>(6.65-18.92) | 16.39<br>(6.35-42.26) | 0.49 | 2.19<br>(0.96-5.02) | 2.5<br>(0.65-9.74) | 0.82 |
| 5 | 17.79<br>(6.98-45.31) | 26.98<br>(7.97-91.28) | 0.44 | 0.53<br>(0.23-1.21) | 0.62<br>(0.136-2.84) | 0.83 |
| 7F | 142.4<br>(25.17-805.4) | 117.4<br>(10.02-1376) | 0.87 | 0.63<br>(0.21-1.86) | 1.20<br>(0.19-7.50) | 0.66 |
| 19A | 23.52<br>(6.64-83.31) | 47.49<br>(5.76-391.3) | 0.63 | 1.11<br>(0.33-3.72) | 2.31<br>(0.4-13.37) | 0.50 |
| 3 | 22.37<br>(8.4-59.54) | 24.31<br>(6.58-89.82) | 0.90 | 0.32<br>(0.15-0.7) | 0.56<br>(0.2-1.63) | 0.48 |

Abbreviations: GMOI (geometric mean opsonophagocytic index), GMC (geometric mean concentration) 95% CI (95% confidence interval), PCV13 (pneumococcal conjugate vaccine 13) The geometric mean opsonic index (GMOI) and 95% confidence intervals (CI) of results are reported for MOPA (HUU n = 11, HEU n = 9) ; the serotype-specific IgG geometric mean concentration (GMC) (HUU n = 11, HEU n = 8) for all PCV13 serotypes (1, 3, 4, 5, 6 A, 6B, 7F, 9V, 14, 18C, 19A, 19F, and 23F) are reported, less than 8 was reported as no response.

### Evidence of HIV exposure in early infancy

Next, we sought to ascertain evidence of HIV exposure as a driver of dysregulation in innate and adaptive immunity of HEU new-borns by measuring HIV specific responses from birth to infancy. We measured HIV Gag-specific responses in peripheral blood mononuclear cells ^67^ using an 18-hour *ex-vivo* IFN-γ ELISpot assay in both the HEU new-born and the longitudinal infant cohorts. (Figure 6a; Supplementary Figure 3a) ^68–70^. Few Gag-specific responses were detectable in cord blood of HEU new-borns (Supplementary Figure 5). Consistent with previous studies ^71, 72^, there was detectable IFN-γ producing HIV Gag-specific cell responses in approximately 50% of the longitudinal cohort of HEU infants (Figure 6b).

**Figure 6:**
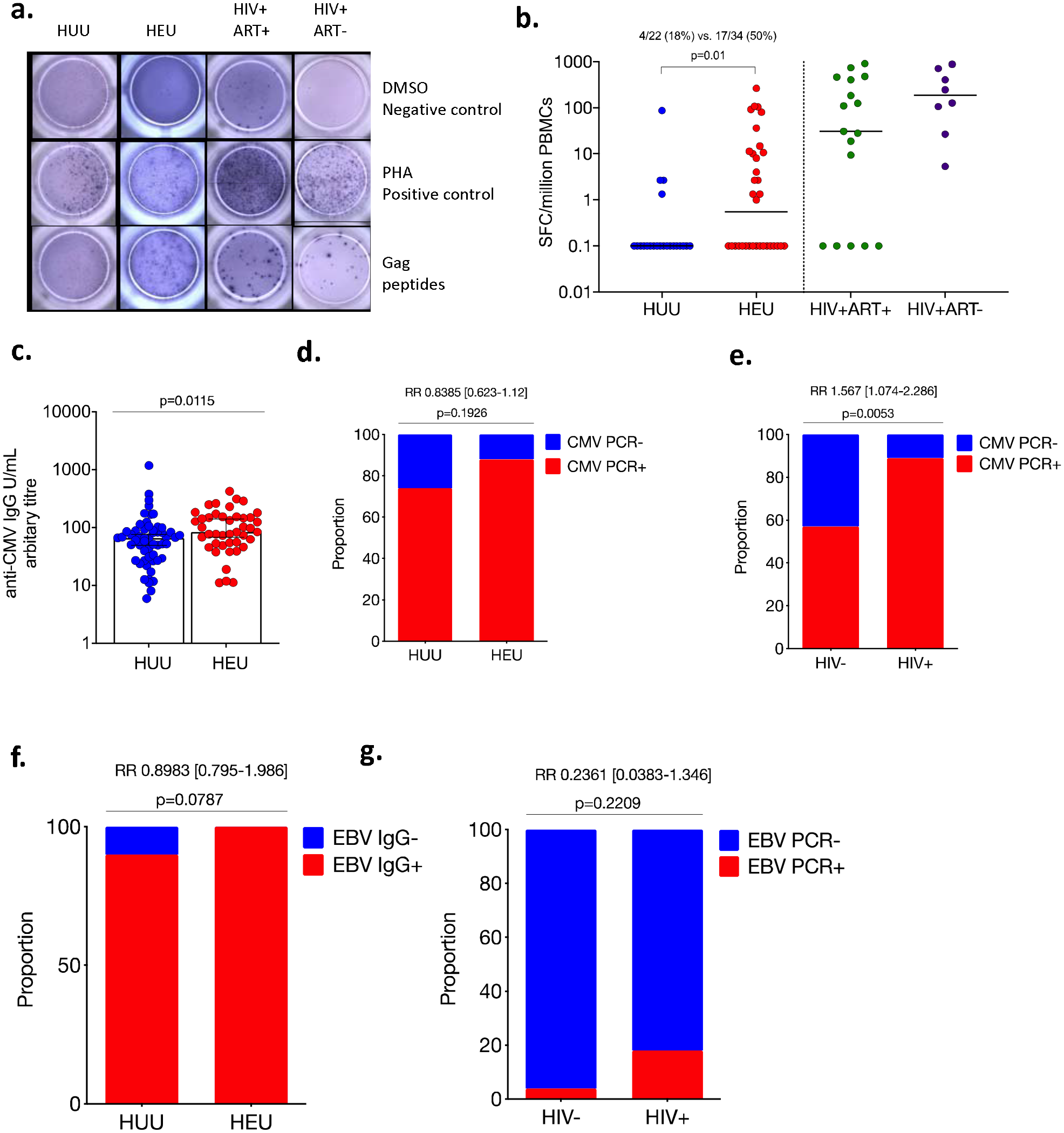
Infant exposure to immune modulating viruses HCMV and HIV. **a)** Detection of Gag-specific T-cells by T-cell ELISPOT assay in HUU and HEU infants, and ART naïve/experienced HIV-infected adults. Isolated PBMCs were incubated with 15-mer Gag peptide pool, PHA as a positive control or RPMI media as a negative control. IFN-γ producing cells were detected on a 96-well microtitre ELISpot plate. **b)** The frequency of SFCs/million PBMCs are plotted for all subjects. Data analysed using Fisher’s exact test (HUU, n =22; HEU, n=34; HIV+ART+, n=17; HIV+ART-, n=8). **c)** Plasma anti-CMV IgG was measured in infants at 5-15weeks of age and assigned an arbitrary titer; blue circles are HUU controls and red circles are HEU. Data are presented as medians [IQR] and analysed using Mann Whitney U test (HUU n=57, HEU n=42). The proportion of **d)** HUU and HEU infants and **e)** HIV-infected and uninfected mothers with RT-PCR detected CMV DNA in their oropharangeal throat swab and breast milk respectively, (HIV-n=23, HIV+ n=35). The proportion of **f)** HUU and HEU infants who were seropositive for anti-EBV IgG in their plasma (HUU n=57, HEU n=42) and **g)** mothers who had detectable EBNA4 protein by RT-PCR, (HIV-n=23, HIV+ n=35). Data analysed using Fisher’s exact test and effective size reported as a relative risk.

### HEU infants have increased exposure to maternal-derived hCMV

HEU infants are more frequently exposed to human herpes viruses (HHV) *in utero* and/or perinatally than HUU infants ^73, 74^, due to the high seroprevalence and reactivation of hCMV and EBV in HIV-infected individuals and pregnant women ^75–80^. We therefore sought to determine the extent of immune-modulating HHV, hCMV and EBV infection in our longitudinal infant cohort.

We found that anti-hCMV IgG titers were higher in sera from HEU infants than HUU controls (Figure 6c), but hCMV PCR detection was similar (Figure 6d), indicating potential differences in the pattern of exposure to hCMV but not increased infection. We then measured hCMV in maternal breastmilk as a potential source of transmission or transfer of viral products to the infants. We found that the proportion of mothers with an hCMV positive PCR result in breast milk was higher in the HIV-infected mothers compared to HIV-uninfected controls (Figure 6e). In contrast, we did not observe any statistically significant differences between potential EBV exposure in HEU infants compared to HUU controls, whether by EBV IgG ELISA (Figure 6f) or PCR (Figure 6g). Overall, these results demonstrate that HHV (hCMV and EBV) infection are common in infancy, irrespective of HIV-exposure status. High hCMV titres in HEU infants may indicate immune boosting from breastmilk of HIV-infected women.

## Discussion

HIV-exposed but uninfected infants are at an increased risk of a wide variety of infections even in the era of universal access to maternal ART, but the underlying immunological basis is not well understood ^7, 51^. We show altered monocyte phagosomal function, dysregulated B cell homeostasis and selective impairment of vaccine responses in HEU infants within the first 6 months of life. We also demonstrate evidence of HIV exposure and increased likelihood of hCMV exposure in HEU infants. HIV and hCMV are immunomodulatory viruses ^81^, so co-exposure to these two pathogens in early life may contribute to this immune dysregulation. We postulate that the complexity, variable severity and/or persistence of this immunological phenotype explain the variable clinical manifestations reported in HEU infants ^8, 9, 11, 40, 82–86^, which may depend on the duration and intensity of exposure to HIV and other infectious co-factors.

The impaired monocyte phagosomal function in HEU new-borns highlights their potential vulnerability to bacterial infection before the primary immunisation series. Monocyte bactericidal activity requires uptake, reactive oxygen species (ROS) formation and phagosomes-lysosomes fusion resulting in inhibition, killing and degradation of internalised bacteria ^87, 88^. In our setting, the validated flow cytometer reporter assay of phagocyte function which uses zymosan has shown poor immune function and superoxide burst activity in HIV-infected adults with active tuberculosis (TB)^89^. Monocyte functional impairment against encapsulated bacteria has also been observed in “age-associated” inflammation, where monocyte-activating cytokines TNF-α and IL-6 are augmented ^90^. Similarly, increased monocyte inflammatory markers, including sTNF-RI, IL-6, IP-10, oxLDL and sCD14 are reported in HEU new-borns ^87^; as well as enhanced pro-inflammatory cytokine secretion following stimulation with diverse PAMPs at 6 weeks of age ^91^. Moreover, recent PBMC transcriptomic profiling in HEU infants at 1-2 years of age, revealed down-regulated genes (LCN2, CAMP, HP, MMP8, BPI, LTF) associated with neutrophil function^92^. It is therefore plausible that the observed impaired monocyte phagosomal function is a consequence of an inflammatory microenvironment in HEU infants, from an inflammatory maternal cytokine milleau and/or exposure to HIV antigens that could explain their increased susceptibility to bacterial infection including pneumonia^14^.

In line with the proposed inflammatory environment in HEU new-borns^93^, we observed high proportions of FcLR4 expressing B cells. In HIV-infected adults, B cells expressing the inhibitory receptor FcLR4 are over-represented, an ‘exhausted’ B cell phenotype, that displays poor B cell receptor mediated activation and antigen-specific antibody production in chronic HIV infection ^60, 94, 95^. Additionally, in HEU infants at 6 to 14 weeks of age, we observed low proportions of tissue-like memory (TLM) and immature-transitional B-cell subsets. TLM and immature transitional B cells are augmented in HIV-infected adults, occurring most profoundly during chronic HIV infection ^96, 97^. The contraction of atypical memory B cells in HEU infants may be indicative of dysregulated B cell homeostasis ^98^. Importantly, our data presents evidence of increased exposure to HIV and hCMV in HEU infants from maternal HIV and/or viral proteins, hCMV recrudescence and high PPD specific IFNγ, that may promote B cell dysregulation in the first 2 months of life. PPD responses are shown to be increased in *mycobacterium tuberculosis* (Mtb) sensitised mothers, which may explain our findings^99^, furthermore, others have described a bimodal response to BCG/PPD (high/low) in HEU infants in our setting^100^. Taken together, the mechanisms of B cell dysregulation are likely distinct from those seen in chronic HIV infection (hypergammaglobulinaemia)^101^, due to the lack of replicative virus and preserved CD4 T-cells.

Altered B cell homeostasis is associated with impaired antibody responses during chronic HIV infection ^102–105^. Consistent with this observation, HIV-infected mothers in our cohort exhibited lower anti-Hib and anti-TT antibody titers than HIV-uninfected mothers. In agreement with maternal titers, we observed low anti-Hib titers in HEU infants. It is known that HIV-infected children are at greater risk of vaccine failure compared to HIV-uninfected children ^106–108^. However, our observation is in contrast with studies conducted in South Africa ^109^ and Uganda ^110, 111^, who reported robust anti-Hib and anti-DT antibody titers in HEU infants. In our setting the Pentavalent Vaccine (DPT-HepB-Hib) was used, in South Africa it was (DTP-Hib and DTaP-IPV/Hib) ^109^, and in Uganda (aP-HibCV and DTwP/HBV/PRP-T) ^110, 111^. Differential vaccine immunogenicity is likely multifactorial, influenced by persistent immune exposure to HIV proteins, the time to maternal ART use and the unique burden of infectious co-factors that likely contribute to a microenvironment of pro-inflammation. Furthermore, HIV proteins are reported to cause aberrant binding to surface immunoglobulins ^112–115^, which may interfere with specific antibody responses. Consistent with previous observations ^116^, we observed that a relatively large number of HEU infants mounted an IFN-γ response following stimulation with HIV Gag, however we did not detect IFN-γ responses in HEU new-borns. Detection of HIV-specific T cell responses in HEU infants was first described nearly 30 years ago ^71, 72^. Others have reported that HEU infants have increased regulatory T cells (Tregs) that correlate with decreased T cell function ^29^ and have shown that depletion of Treg cells from the cord blood of HEU new-borns reveals HIV-1-specific T responses ^116^. We also observed poor IFN-γ responses in cord blood of HEU new-borns following stimulation with the T cell mitogen PHA (which crosslinks the TCR and glycosylated surface proteins). Taken together, these data point towards HIV exposure as a possible driver of increased regulation/inhibition of T cell activation at birth.

Our study has some limitations, including use of a surrogate assay to measure monocyte function and the limited number of assays we could perform on each sample. Additionally, our cohorts were too small to establish direct associations with immune dysregulation and clinical presentation of disease.

In conclusion, we show altered monocyte phagosomal function, dysregulated B cell homeostasis and selective impairment of vaccine antibody responses in HEU infants within the first 6 months of life. This period of vulnerability likely contributes to increased susceptibility to disease-causing bacteria that commonly cause life-threating illness such as pneumonia in HEU infants.

## Methods

### Study Design and population

The study was conducted in Southern Malawi, at Ndirande Health Centre (NHC) (a primary healthcare facility in the city of Blantyre) and at Queen Elizabeth Central Hospital (QECH) (a tertiary teaching hospital in the city of Blantyre). We recruited the infant cohort in two contiguous groups that were followed longitudinally. The first group were aged 5-9 weeks (pre-1^st^ vaccine dose) who were followed up to age 14-15 weeks (post-2^nd^ vaccine dose) and the second group were aged 14-15 weeks (post-2^nd^ vaccine dose) who were followed up to 18-23 weeks of age (post-3^rd^ vaccine dose). We recruited pregnant women in the early stages of labour at QECH maternity ward and subsequently their babies at birth (termed birth cohort). Participating mothers were healthy, asymptomatic adults (>18 yr) comprising HIV-infected and HIV-uninfected volunteers. HIV testing was performed on whole blood using two commercial point-of-care rapid HIV test kits, Determine HIV 1/2 kit (Abbott Diagnostic Division, Abbott Park, IL) and Unigold HIV 1/2 kit (Trinity Biotech Inc., Bray, Ireland). HIV-infected participants received first line ART (Option B+ (Lamivudine, Tenofovir DF and Efavirenz (3TC/TDF/EFV)) at any point during pregnancy and had a CD4+ T-cell count performed. All HEU babies received single dose nevirapine at birth and co-trimoxazole until 6 months of age. Exclusion criteria for the study participants were current or past history of smoking, heart disease, tuberculosis, high blood pressure, drug use, syphilis, severe anaemia (haemoglobin <8 g/dl), placental abnormalities, infant death and existing comorbidities. All babies received Bacille Calmette Guerin (BCG) vaccine at birth. Written informed consent was obtained from all participants prior to recruitment. Ethical approval was obtained from the University of Malawi College of Medicine Research and ethics committee (COMREC), Blantyre, Malawi, protocol numbers P.11/11/1140, P.06/11/1088.

### Sample collection and processing

To maximise the number of samples we were able to collect from very young infants, we collected 5ml of venous blood from the infants, at 5-9, 14-15 and 18-23 weeks of age following attendance at the vaccination clinic for pentavalent DPT-HepB-Hib immunisation. Participants who did not complete their vaccine course were excluded from the analysis. From their mothers, 10ml of venous blood were collected only at the first visit and 3mls of breastmilk at all three time points. We collected upto 40ml cord blood from the umbilical vein into sodium heparinised tubes using a 50ml syringe from the baby interface of the placenta immediately after birth. Whole blood was kept at RT for no longer than 2 hours from collection to processing. Peripheral blood mononuclear cells were isolated by density centrifugation. Diluted whole blood was layered on top of Histopaque Ficoll gradient (Sigma-Aldrich, UK) at a ratio of 2:1 and centrifuged (900g, 24 ᵒC for 30 minutes). The PBMC fraction was removed using a Pasteur pipette and washed immediately in HBSS (Sigma Aldrich, UK) by centrifugation (500g, 4 ᵒC for 5 minutes). Cells were re-suspended in 2mL of complete media ((RPMI 1640 plus (Sigma-Aldrich, UK) with 10mM HEPES (Sigma-Aldrich, UK), 1% v/v penicillin, 1% V/V streptomycin (Sigma Aldrich) and 2mM L-glutamine (Sigma-Aldrich, UK)), then agitated to ensure complete mixing and counted using a haemocytometer with 1:10 trypan blue (Sigma Aldrich, UK) staining to confirm viability. Plasma was separated by centrifugation at 1500rpm for 10 minutes, aspirated, aliquoted and stored at −80 for later use. Breastmilk samples were collected by hand expression, fractionated into lipid and aqueous phase and stored at −80. Due to limitations in the volume of blood collectable from very young babies and limited cell numbers, not all the assays were performed on samples obtained from every participant.

### HIV testing

New-borns and infants qualitative HIV DNA PCR tests were performed in batches of 23. Total RNA was isolated from 0.5 x10^6^ cells using AMPLICOR HIV-1 DNA test, V1.5 (Roche, USA) according to the manufacturer’s instructions.

Three HIV-RDT kits, from two separate manufacturers (Unigold™ and Determine™) were used to confirm the presence or absence of HIV specific immunoglobulins (Ig) in maternal peripheral blood. Cord blood mononuclear cells (CBMCs) from were stored in 500ul of RNAlater and analysed at UCL by digital PCR as described elsewhere ^117^.

### Maternal CD4 counts

Peripheral blood CD4 T-counts and full blood count (FBC) were performed at the Malawi-Liverpool Wellcome Trust Clinical Research Programme Diagnostic Laboratory on an Hmx analyser (Beckman Coulter, USA) using a standardized protocol.

### Phenotypic analysis

Multicolour flow cytometry analysis was performed on whole blood (WB). Samples were stained with the following fluorochrome conjugated antibodies, anti-CD14 Phyocoerythrin Cyanine-7 (PECy7), anti-CD3 Allophycocyanin-H7 (APC-H7), anti-CD4 Pacific Blue (PB), anti-CD8 Fluorescein (FITC), anti-CD8 (PECY7), CD45RA Phyocoerythrin (PE), CD45RA PECY5, CCR7 Allophycocyanin (APC), anti-CD19 (APC), anti-CD19 Peridinin-Chlorophyll-Protein (PERCP), anti-CD27 (PE), CD10 PE-Cy7, CD21 FITC, CD27 APC-Cy7, CD95 e450, FcLR4 PE, CD57 FITC, PD-1 APC, PD-1 PE (Supplementary Tables 1 and 2, Gating strategy supplementary figure 2). Samples were acquired on Beckman Coulter Cyan ADP and analyses were performed using FlowJo Version 7.6.5 and 10.5 software (Treestar).

### Measurement of monocyte phagosomal enzymatic activity

Phagosomal oxidative burst and bulk proteolytic function in monocytes was measured using a flow cytometry–based reporter bead assay as described previously^57, 118, 119^. Briefly, 3μm diameter silica beads were derivatized with the calibration fluorochrome (Alexa Fluor 405-SE) and the fluorogenic reporter substrates Oxyburst Green succinimidyl ester (Oxybeads) (Molecular Probes, Eugene, OR) for superoxide burst, or DQ Green bovine serum albumin (DQ-beads) (Molecular Probes, Eugene, OR) for bulk proteolysis. When the beads are internalised by monocytes the fluorescence intensity is proportional to the degree of activity in the phagosome, which provided a readout of phagosomal enzymatic activity. The readout for the assay is reported as the ratio of the median fluorescent intensity of the reporter fluorochrome at 60mins:10mins and 240mins:10mins for oxidative burst and bulk proteolysis, respectively. The uptake cut-off was 35% in the assay.

### T cell IFNγ ELISpot

2×10^6^ PBMCs were stimulated in an 18-hour IFN-γ T cell ELISpot; plates were coated with purified anti-human interferon-γ monoclonal antibody (αhuman-IFN-γ-mAb) 1-D1K (Mabtech, UK) 1:66 at a concentration of 3.78µg/mL in sterile PBS. The enumerated IFN-γ producing cells after antigen stimulation were used as an index of effector memory T cell responses, as previously described ^120^. Data were reported after deducting 2x the value of the negative well. Stimulating antigens were: Phaseolus vulgaris lectin (PHA) 5ug/ml (NIBSC, UK), purified protein derivative (PPD) 10ug/ml (Statens serum institute, Denmark) and HIV-1 consensus C gag 15-mer peptides (GAG peptide) 10ug/ml (NIH AIDS, USA)^120^

### Human cytomegalovirus PCR

Real time PCR was used to detect hCMV in HIV-infected and uninfected maternal breast milk and plasma. The results were based on a standard curve constructed using an in-house plasmid previously calibrated against quality controls for molecular diagnostics and external quality assessment panels (Hamprecht et al. 1998). The positive control was assessed again and an expected value of 2,500 copies seen with results read on the ABI Prism 7500 (Thermofisher, UK). The amplification plot was assessed against a standard slope range of −3.10 to −3.60 and a R^2^ value > 0.9.

### Detection of Cytomegalovirus specific IgG and IgM antibodies

hCMV specific IgM in plasma was measured using a commercial ELISA kit (IBL International, Hamburg) according to manufacturer’s instructions. A semi-quantitative, in-house hCMV IgG assay was used at the laboratories in the University of Birmingham, UK. Plasma was diluted 1/4 in a dilution in a 1:1:1 mix of plasma from three healthy donors, the top concentration was assigned an arbitrary unit of 1000. The unknown samples were related to the standard curve and a titre calculated. Briefly, Nunc 96-well plates Maxisorb (Fisher Scientific # 442404) were coated with 50μL UV activated hCMV-lysate (1:4000) and mock-lysate (1:4000) in carbonate-bicarbonate buffer pH 9.6 (sigma capsules), then covered with parafilm and incubated at 4°C overnight in the fridge. The following day, plates were washed three times in 200μL (PBS + 0.05% Tween20), 100μL of samples were added 1:600 in dilution buffer (PBS + 1%BSA + 0.05% Tween20), blank dilution buffer, and standards, then incubated for 1h at room temperature (RT). Plates were washed again three times, then 100μL of the secondary antibody, anti-human IgG-HRP (1/8000 dilution in PBS+1% BSA+0.05% Tween20) goat anti-human IgG (Southern Biotech #2040-05). Plates were incubated and washed again as described, after which 100μL of tetramethyl benzidine (TMB-solution) (Tebu-Bio) was added for 10 min at RT. The reaction was stopped with 100μL 1M HCl and plates were read at 405nm (Biotek instrument).

### Sandwich enzyme-linked immunosorbent assay to detect IgG specific to vaccine antigens

In an in-house ELISA, TT or DT (both NIBSC, UK) were diluted to 0.5 Lf/mL or Hib capsular polysaccharide 0.1μg/mL in 10 mL carbonate coating buffer (0.015 M Na_2_CO_3_, 0.035 M NaHCO_3_ pH9.6); 100 μL per well of the solution was pipetted into a 96-well flat bottom Maxisorp plate. Plates were incubated overnight at 4 °C then washed seven times with PBS 0.05% Tween. 50 μl goat anti-human alkaline phosphatase-conjugated secondary antibodies (Southern Biotech, UK) were diluted to 1ug/mL in PBS 0.05% Tween 2% BSA and added to each well. The plates were incubated for 1 h at 37°C, then washed seven times with PBS 0.05% Tween. 100 μL of Sigma-fast p-nitrophenyl phosphate substrate was added to each well (Sigma, UK). A standard curve was generated using a set of 2-fold dilutions of a standard pooled serum (NIBSC, UK). Optical density was measured (without acid stopping the reaction) after 10 min using an ELISA plate reader (Biotek, UK) set at 405nm and SoftMax Pro software ^121^.

### Multiplexed opsonophagocytosis killing assay and serotype-specific IgG

Sera from 20 infants were sent on dry ice to the UCL Great Ormond Street Institute of Child Health (UCL; United Kingdom). Infants had received 3 doses of Prevnar (PCV13) at 6, 10, and 14 weeks of age. Sera were stored at –20°C until analysis. Analyses were performed at the World Health Organization pneumococcal reference laboratory (University College London, United Kingdom). Immunoglobulin G (IgG) serum concentrations specific for the 13 vaccine serotypes (1, 3, 4, 5, 6 A, 6B, 7F, 9V, 14, 18C, 19A, 19F, and 23F) were measured using an enzyme-linked immunosorbent assay (ELISA) after adsorption with cell-wall and 22F polysaccharides to increase the assay specificity ^122^. A standardized opsonophagocytic assay (OPA) was used to measure functional antibodies against the same serotypes ^123^. The OPA titre was defined as the reciprocal of the lowest serum dilution that induces ≥50% bacterial cell death compared to the assay control.

### Statistical analysis

Statistical analysis and graphical presentation were performed using Prism 7/8 (GraphPad Software, San Diego, USA) and Python (Python Software Foundation) was used to calculate summary statistics. Demographic and clinical characteristics were compared using Mann-Whitney-U for continuous and Fisher’s exact tests or χ^2^ for discrete variables. ELISpot data were reported as subtracted 2 x background. Serotype-specific opsonophagocytic indexes (OIs) were reported using geometric means and 95% confidence intervals. The OIs were classified as being positive or negative based on the current recommended cut-off value of <8 (negative) and ≥8 (positive). Results are reported as median and IQR as stated.

## Data Availability

The data is not available.

## Acknowledgments

The authors are grateful to the participants for their willingness to participate in this study. This work was supported by a studentship from the Franklin Adams Trust and a project grant from the Wellcome Trust. The Malawi-Liverpool-Wellcome Trust Clinical Research Programme was supported by a strategic award from the Wellcome Trust.

## Author contribution

L.A. A.F, R.S.H, S.R.J, D.M contributed to study design. L.A, A.B, B.F, R.K, D.H.B, D.G, P.M, D.M performed the experiments. W.N recruited and monitored the participants. L.A, K.C.J analysis and interpretation. H.M, D.M, A.F., K.C.J, R.S.H, reviewing the manuscript L.A, K.C.J, S.R.J, A.F, R.S.H, final approval.

## Supplementary Figures

**Supplementary Figure 1:**
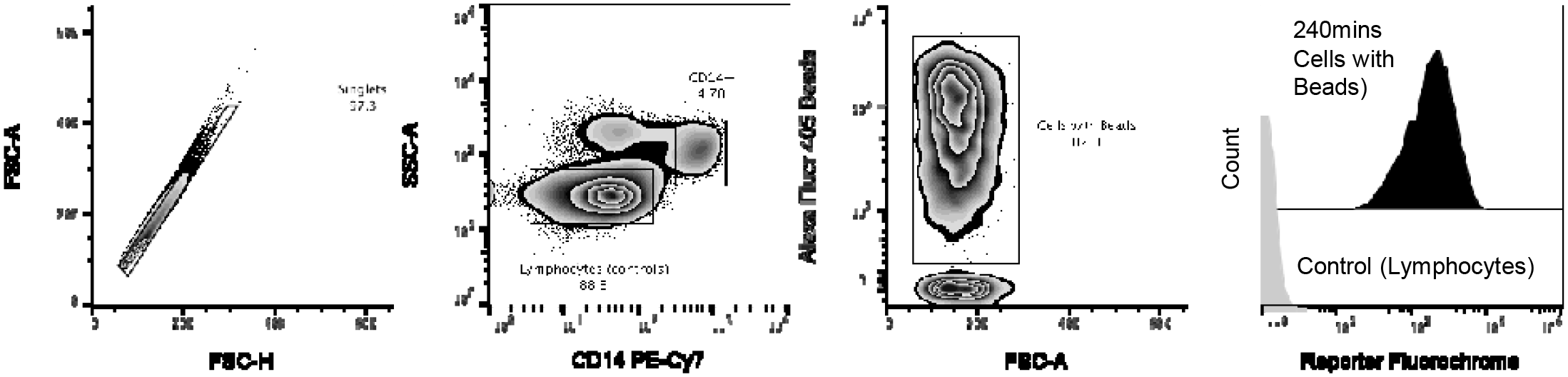
Gating strategy to detect monocyte phagosomal functional capacity at birth. Singlets were defined using FSC-A vs. FSC-H parameters and lymphocytes were gated using SSC-A and CD14 PE-CY7. Then FSC-A vs. AF405 to gate cells with beads. The readout for the assay are reported as the median fluorescent intensity of the reporter fluorochrome at 60mins:10mins and 240mins:10mins for oxidative burst and bulk proteolysis, respectively. The activity index was calculated using a ratio of the reporter fluorochrome over the the calibration fluorochrome. Only individuals with an uptake of greater than ≥30% were used in the phagosomal analysis.

**Supplementary Figure 2:**
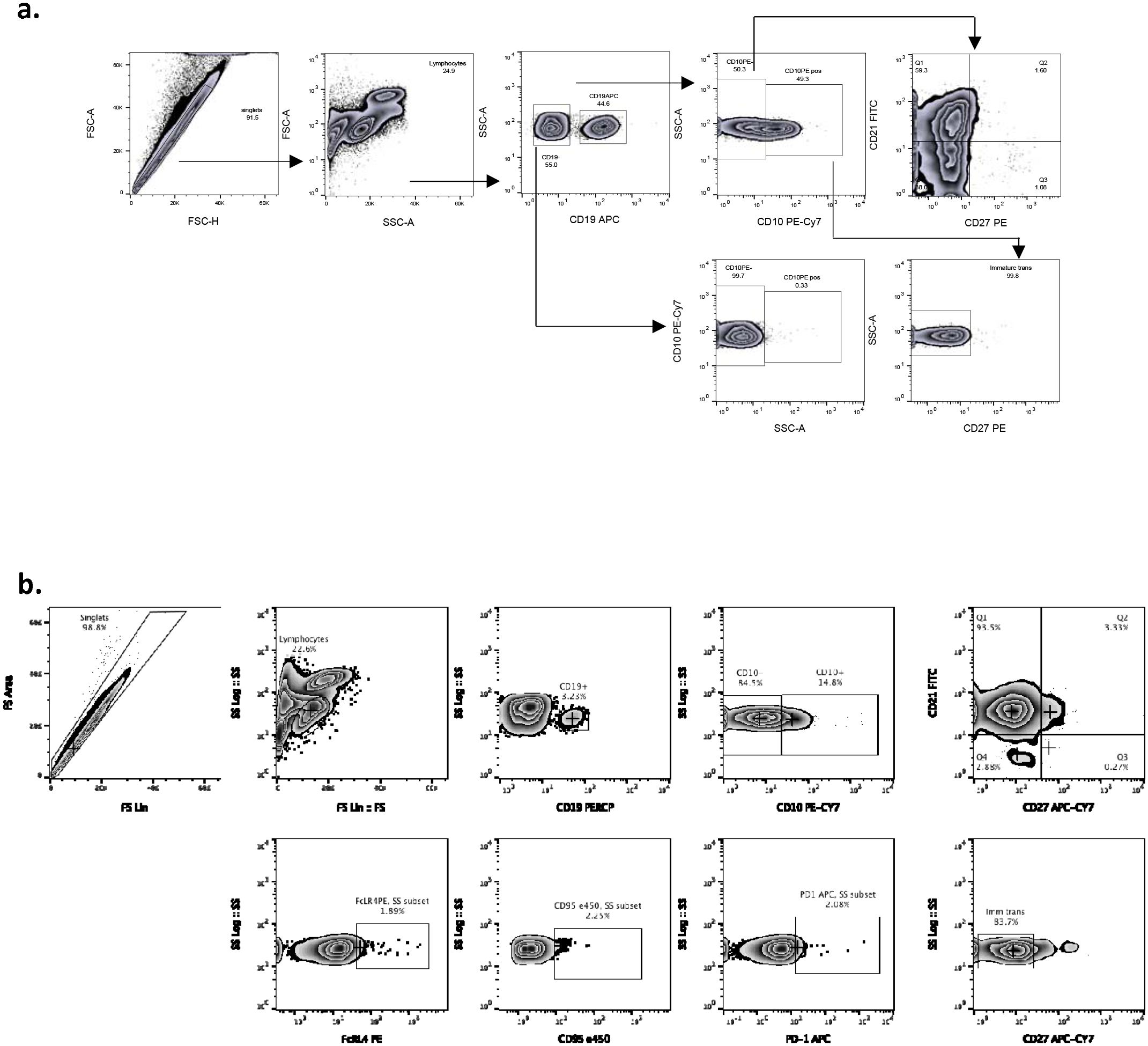
Gating strategy to characterise B cells. **a)** Blood was stained with the following fluorochrome-conjugated antibodies, anti-CD19 APC, anti-CD10 PE-Cy7, anti-CD21-FITC and anti-CD27 PE. Singlets were defined using FSC-A vs. FSC-H parameters and lymphocytes were gated using SSC-A and FSC-A. B cells were then gated using CD19 against SSC-A. The following populations were determined: CD10^-^ CD21^+^CD27^-^ (naive), CD10^-^CD21^-^CD27^+^ (resting memory), CD10^-^CD21^+^CD27^+^ (activated memory), CD10^-^CD21^-^CD27^-^ (tissue-like memory) and **c)** CD10^+^CD27^-^ (immature transitional). **b)** Blood was stained with CD19 PERCP, anti-CD10 PE-Cy7, anti-CD21-FITC and anti-CD27 APCCY7, FcRL4 PE, CD95e450 and CD27 APCCY7 to determine B cells expressing inhibitory markers.

**Supplementary Figure 3:**
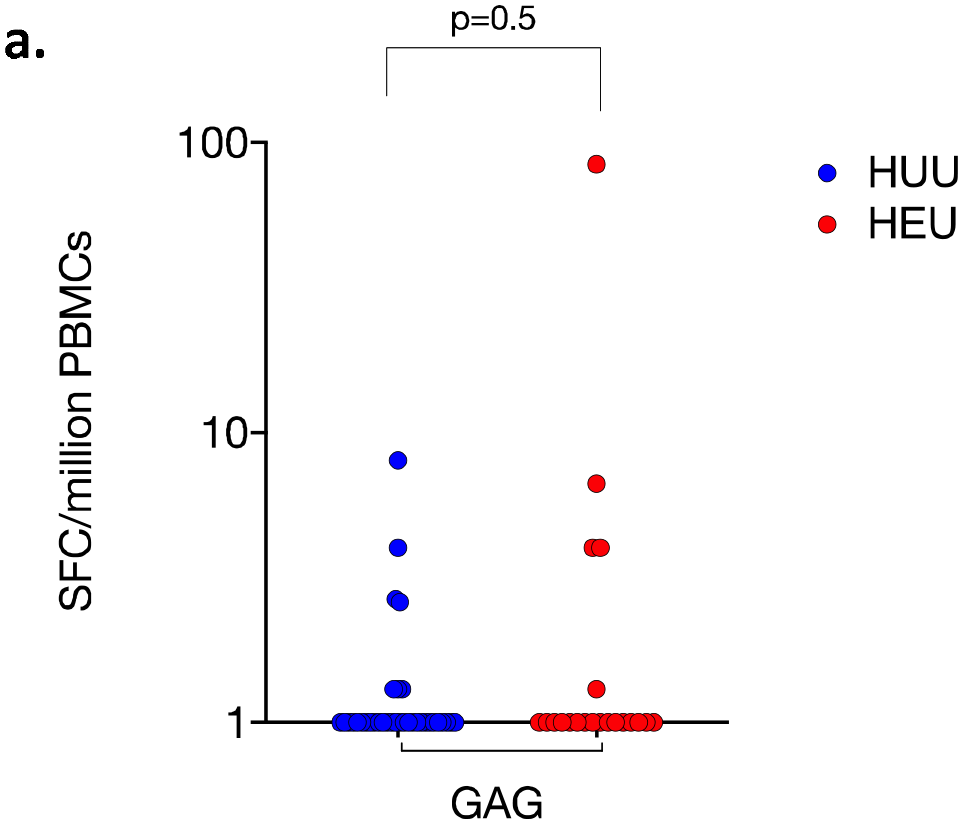
IFN-γ producing cells to HIV protein GAG at birth. A 15-mer GAG peptide pool was used to stimulate isolated CBMCs from cord blood in an 18hour T cell EliSpot assay. IFN-γ producing cells were detected on a 96-well microtitre ELISpot plate. **a)** The frequency of GAG SFCs/million CBMCs are plotted for all subjects. Data analysed using Fisher’s exact test (HUU, n =35; HEU, n=23).

**Supplementary Figure 4:**
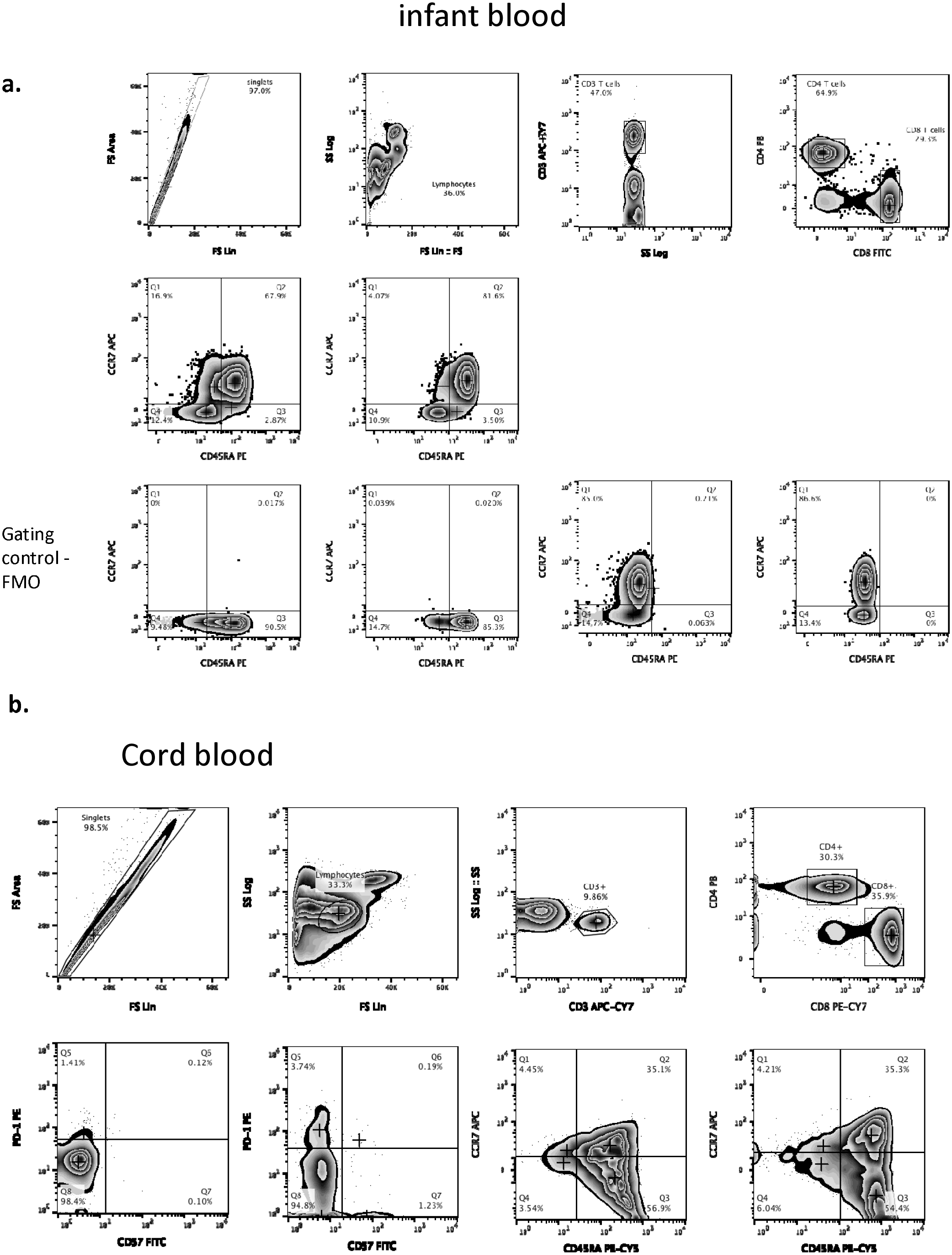
Gating strategy to characterise T cell subsets. **a)** Whole blood was stained with the following fluorochrome-conjugated antibodies, anti-CD3 APCH7, anti-CD4 Pacific Blue, anti-CD8-FITC, anti-CCR7 APC and anti-CD45RA PE. **b)** Cord blood was stained with the following fluorochrome-conjugated antibodies, anti-CD3 APCY7, anti-CD4 Pacific Blue, anti-CD8-PECY7, anti-CCR7 APC and anti-CD45RA PECY5, PD-1 PE and CD57 FITC. Singlets were defined using FSC-A vs. FSC-H parameters and lymphocytes were gated using SSC-A and FSC-A. T cells were then gated using CD3 against SSC-A, then a CD4 versus CD8 plot was used to separate the two main T cell subsets. FMO were used to determine gating. CD4^+^ and CD8^+^ T cell subsets were clasified using CCR7 and CD45RA as follows CCR7-CD45RA-(effector memory), CCR7^+^CD45RA-(central memory), CCR7^+^CD45RA^+^ (naïve) and CCR7^-^CD45RA^+^(terminally-differentiated).

**Supplementary Figure 5:**
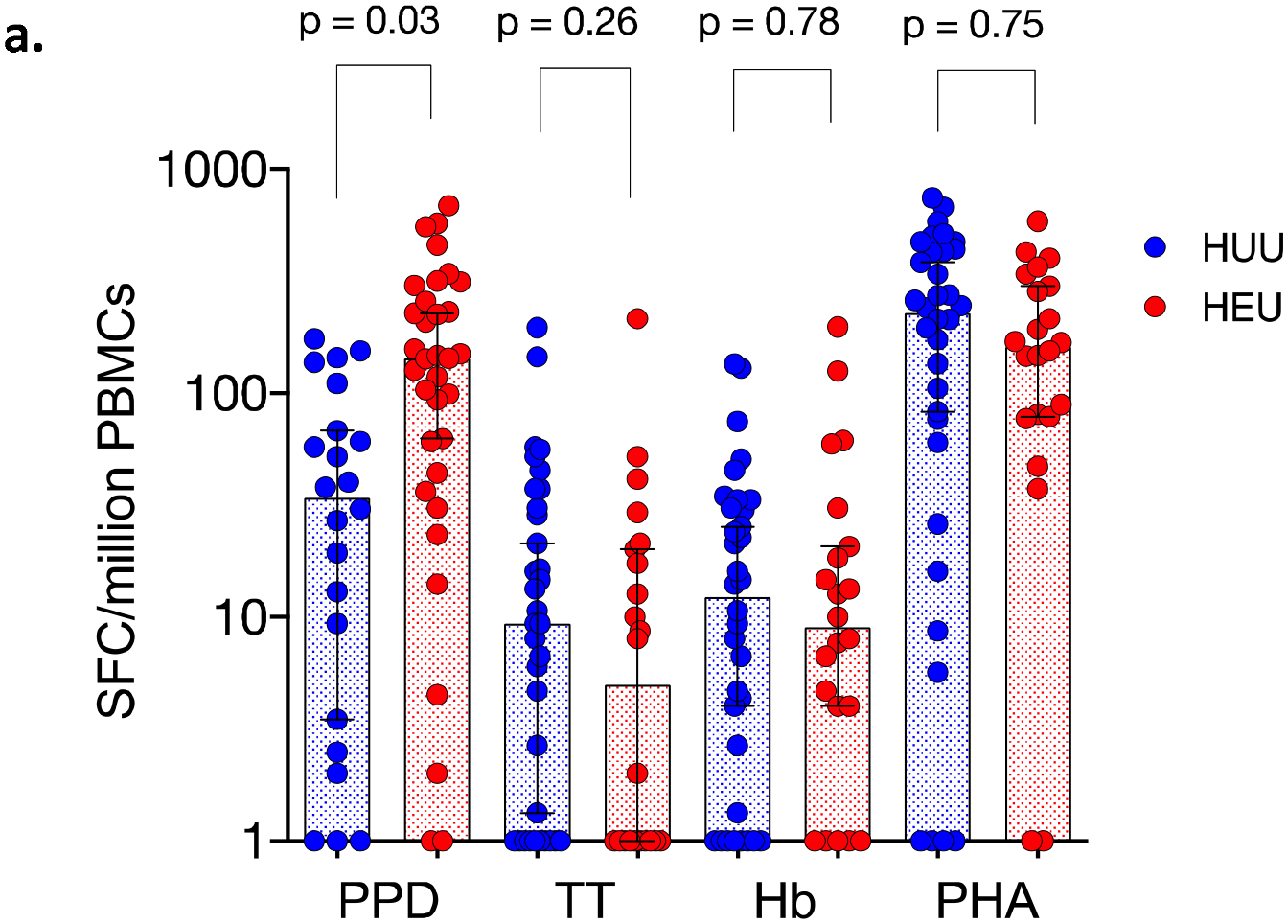
IFN-γ producing cells to vaccine antigens in HEU and HUU infants. Detection of IFN-γ producing cells using a T-cell ELISPOT assay in HUU and HEU infants.. Isolated PBMCs were incubated with either PPD, TT, Hb or PHA as a positive control or RPMI media as a negative control for 18 hours. IFN-γ producing cells were detected on a 96-well microtitre ELISpot plate. **a)** The frequency of SFCs/million PBMCs are plotted for all subjects. Data analysed using Fisher’s exact test (HUU, n =22; HEU, n=34).

## Supplementary Table

**Supplementary Table 1.**
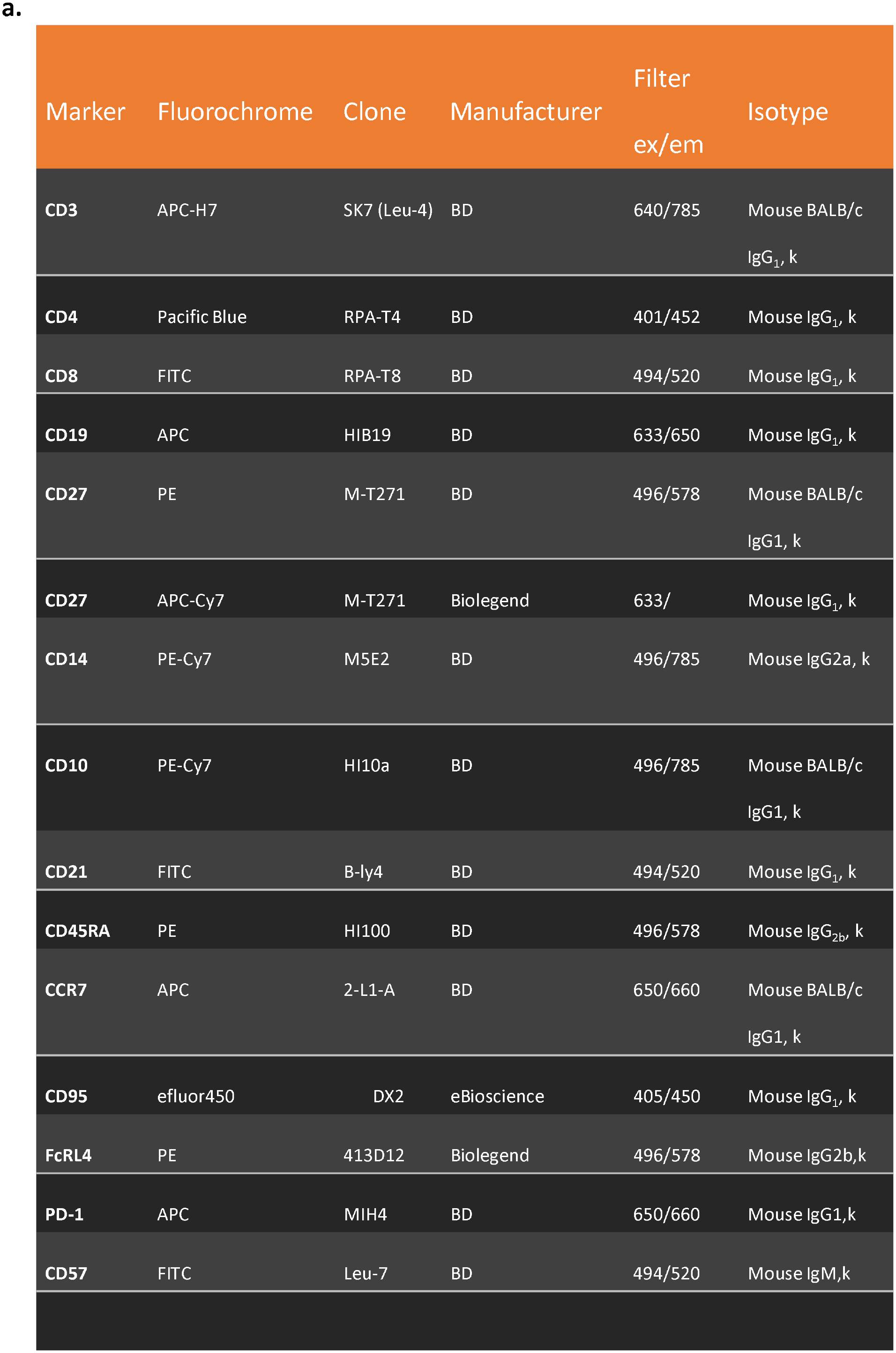
Fluorochromes.

**Supplementary Table 2.** Fluorescent antibody panels.

a.
| Panel 1 | Panel 2 | Panel 3 | Panel 4 | Panel 5 |
| --- | --- | --- | --- | --- |
| anti-CD14<br>Phycoerythrin<br>Cyanine-7<br>(PECy7) | anti-CD19<br>Allophycocyanin (APC) | anti-CD19<br>Peridinin-Chlorophyll-Protein (PERCP) | anti-CD3<br>Allophycocyanin-H7 (APC-H7) | anti-CD3<br>Allophycocyanin-H7 (APC-H7) |
| anti-AF405<br>Beads | anti-CD10 PE-Cy7 | anti-CD10 PE-Cy7 | anti-CD4 Pacific<br>Blue (PB) | anti-CD4 Pacific<br>Blue (PB) |
|  | anti-CD21 FITC | anti-CD21 FITC | anti-CD8<br>Fluorescein<br>(FITC) | anti-CD8 (PECY7) |
|  | anti-CD27 (PE) | anti-CD27 APC-Cy7 | anti-CD45RA<br>Phycoerythrin<br>(PE) | anti-CD45RA<br>PECY5 |
|  |  | anti-CD95 e450 | anti-CCR7<br>Allophycocyanin<br>(APC) | anti-CD57 FITC |
|  |  | anti-FcLR4 PE |  | anti-PD-1 PE |
|  |  | anti-PD -1 APC |  |  |

